# Comparison of Imputation Strategies for Incomplete Electronic Health Data

**DOI:** 10.1101/2025.08.01.25332573

**Authors:** Shuo Zhang, Zhilong Zhang, Yuxi Zhou, Shenda Hong, Huixin Liu

## Abstract

Missing data is a persistent challenge in electronic health records (EHRs), often compromising data integrity and limiting the effectiveness of predictive models in healthcare. This study systematically evaluates five widely used imputation strategies—GAIN, MICE, Median, MissForest, and MIWAE—across three real-world clinical datasets under varying missingness mechanisms (MCAR, MAR, and MNAR) and missingness rates (10%–90%). We assessed imputation quality using multiple statistical measures and examined the relationship between imputation accuracy and downstream classification performance. Our results show that MICE and MissForest consistently outperform other methods across most scenarios, while deep learning-based approaches such as GAIN exhibit high instability under MAR and MNAR, particularly at higher missingness levels. Furthermore, imputation quality does not always align with classification performance, underscoring the need to consider task-specific goals when selecting imputation strategies. We also provide a practical framework summarizing method recommendations based on missingness type and rate, aiming to support robust data preprocessing decisions in clinical AI applications.

## Introduction

Missing data is a pervasive issue in electronic health records (EHRs) and poses a major challenge for clinical research and digital health applications^1,2^. Such missingness can arise from various sources, including incomplete data collection, privacy constraints, heterogeneous data integration, and selective clinical testing. These factors contribute to complex and often non-random missing data patterns. According to the widely accepted taxonomy, missingness can occur under three mechanisms: missing completely at random (MCAR), missing at random (MAR), and missing not at random (MNAR)^3^. In clinical datasets, true MCAR is rare; missingness is more frequently driven by observed or unobserved covariates, resulting in MAR or MNAR mechanisms^4,5^. For instance, in intensive care settings, critically ill patients may be unable to undergo certain tests, leading to systematic missingness in key clinical variables, while less severe cases are more fully documented. These patterns are often correlated with disease severity, and inappropriate imputation strategies may introduce bias, ultimately compromising model performance and clinical decision-making.

To address this issue, a variety of imputation strategies have been proposed^5^. Traditional methods, such as mean or median imputation, multiple imputation by chained equations (MICE^6^), and MissForest^7^, have shown strong performance under relatively simple missingness patterns, especially MCAR. More recently, deep learning-based approaches—such as Generative Adversarial Imputation Networks (GAIN)^8^ and methods based on Variational Autoencoders (MIWAE)^9^—have been introduced to capture more complex structures. While these techniques offer theoretical advantages, their robustness and generalizability in real-world clinical scenarios remain underexplored.

Previous studies have primarily emphasized imputation accuracy, often neglecting the potential distributional shifts induced by imputation and their downstream impact on predictive performance. Moreover, many evaluations are conducted under the MCAR assumption, providing limited insight into the more realistic MAR and MNAR mechanisms. There is still no clear consensus on which imputation methods perform best under different types and levels of missingness, nor on how imputation quality translates into performance in clinical prediction tasks^10^. There is also little guidance on thresholds beyond which imputation may no longer be practically useful.

To fill these gaps, we systematically evaluate five widely used imputation methods—mean/median imputation, MICE, MissForest, GAIN, and MIWAE—under three missingness mechanisms (MCAR, MAR, MNAR) and a range of missing rates, across multiple real-world clinical datasets. We assess both imputation quality and downstream classification performance using multiple metrics. Our goal is to provide practical guidance for selecting appropriate imputation strategies in digital health research and to support the development of more robust and interpretable clinical prediction models.

## METHODS

### Dataset

In this study, we focus on three real-world clinical datasets: MIMIC-III, Breast Cancer, and Kaggle COVID-19. The MIMIC-III dataset is a complete dataset (i.e., no missing values). The Breast Cancer and Kaggle COVID-19 datasets exhibit their inherent missingness. Data usage for this study was approved by the data providers.

MIMIC-III is a publicly available critical care database containing de-identified clinical data from patients admitted to the intensive care units (ICUs) of Beth Israel Deaconess Medical Center between 2001 and 2012^11^. For this study, we extracted a subset comprising 7,214 unique patients with 14 numerical clinical features, along with corresponding survival outcomes. As this dataset is fully observed, it enables the generation of simulated data with varying missing mechanisms and rates.

The Breast Cancer dataset originates from oncology data collected at Memorial Sloan Kettering Cancer Center (MSKCC) between April 2014 and March 2017^12^. It includes genomic analysis data for 1,918 tumor samples, with detailed clinical variables, treatment information, and outcomes for 1,756 patients. For this study, we used a preprocessed version containing 16 key features and retained the natural missingness in the data to more accurately evaluate the performance of different imputation methods on real-world medical data.

The Kaggle COVID-19 dataset is sourced from the COVID-19 Missing Data Project by Cambridge University^13^. This dataset includes clinical records for COVID-19 patients and simulates different missing data patterns to explore the applicability of missing data handling methods for pandemic-related medical data. It is characterized by significant heterogeneity and dynamic changes, making it suitable for validating the generalization capabilities of imputation methods across various types of medical data. The version used in this study contains data from 1,925 patients with 19 key features.

Use of all datasets complied with their respective access and usage guidelines.

### Data Preprocessing

The MIMIC-III dataset consists solely of numerical variables. In contrast, the Breast Cancer and COVID-19 datasets include numerical, categorical (both nominal and ordinal), and multilevel variables. Prior to analysis, categorical variables were transformed using one-hot encoding, while ordinal variables were encoded as integers. This ensured compatibility across imputation and modeling pipelines.

### Outcome Variables

For the MIMIC-III, Breast Cancer, and Kaggle COVID-19 datasets, we used survival status as the outcome variable of interest. Survival status was chosen as the primary outcome to reflect clinically meaningful endpoints across the three datasets, formulated as a binary classification task.

### Missingness Simulation

In this experiment, we simulated missingness in the three datasets mentioned above using three different missing data mechanisms: MCAR, MAR, and MNAR. For MCAR, we custom-built a function to generate missing data. Specifically, we randomly generated a mask matrix matching the dimensions of the data, where the values were determined by a pre-set missing rate to decide whether to assign a missing value (NaN) to each data point. The missing rate varied from 10% to 90% by 10%.

For MAR and MNAR, we use the Pyampute package to automatically generate missing data^14^. By default, both the MAR and MNAR mechanisms randomly selected half of the variables for missingness. In the MAR mechanism, the missingness is related to observed variables but not to the missing values themselves. We chose a subset of features to be missing and used other variables to predict the missingness probability based on the observed features. In the MNAR mechanism, missingness is related to the feature values themselves. To simulate this non-random missingness, we controlled the missingness probability using the feature values, representing situations where severely ill patients could not undergo certain tests due to their condition. This missing pattern is more complex and often occurs in real data collection processes. The missing rate was also set to range from 10% to 90% to evaluate the imputation methods at different missingness levels. These three missing mechanisms allow us to comprehensively simulate various missingness patterns encountered in practical applications and provide a diverse testing environment for evaluating imputation methods.

### Imputation Methods

In this study, we considered the five most popular imputation methods from the literature: Median, MICE (Multiple Imputation by Chained Equations), MissForest, Generative Adversarial Imputation Network (GAIN), and Variational Autoencoder-based MIWAE (Missing Data Imputation with Wasserstein Autoencoders). These methods cover a range from the simplest single imputation techniques to more complex multiple imputation methods, including both traditional and deep learning approaches.

### Classification Model

To assess the utility of the imputed data in predictive tasks, we trained a Random Forest classifier on each completed dataset after imputation. Random Forest is a robust ensemble learning method known for its effectiveness in handling high-dimensional, heterogeneous clinical data and its resistance to overfitting^15–17^. Each experiment was repeated five times per imputation method and missingness level to account for variability due to random seed effects. Results were summarized using mean and standard deviation.

### Evaluation Metrics

In this study, we employed several commonly used evaluation metrics to measure the performance of the imputation methods, with a focus on Root Mean Squared Error (RMSE) and Area Under the Receiver Operating Characteristic Curve (AUC), which are key indicators of imputation performance. RMSE was used to evaluate the difference between imputed data and the original data, with smaller values indicating better imputation quality (i.e., the imputed data is closer to the true data)^18^. AUC was used to evaluate the performance of the imputed data in downstream classification tasks, especially its ability to predict binary classification tasks^19^. Compared to RMSE, AUC places more emphasis on measuring whether the imputed data can provide effective discriminative power for the classification model^20^. Therefore, the comparison between RMSE and AUC helps provide a comprehensive understanding of the relationship between imputation quality and downstream task performance. Additionally, we used other supplementary metrics, such as Mean Absolute Error (MAE), R-squared (R^2^), Kullback-Leibler Divergence (KL divergence), Kolmogorov-Smirnov statistic (KS test), and Wasserstein distance. These metrics offer a more detailed evaluation of imputation quality, particularly in terms of data distribution and feature differences. Furthermore, we ranked the RMSE and AUC for the five imputation methods under different missing rates.

## RESULTS

### Traditional methods achieve lower RMSE and more stable estimation quality

Figure 1 presents the RMSE values of five imputation methods across three datasets—MIMIC-III, Breast Cancer, and COVID-19—under MCAR, MAR, and MNAR mechanisms with missing rates ranging from 10% to 90%. Across all datasets and mechanisms, MICE and MissForest show relatively lower RMSE values in most settings. Their RMSE values tend to increase gradually as the missing rate increases, but the overall variation remains limited. The RMSE values of GAIN are relatively low under the MCAR mechanism when the missing rate is below 50%, but increase more rapidly at higher missing rates. Under MAR and MNAR, GAIN shows more fluctuation. MIWAE also exhibits fluctuations in RMSE under different mechanisms and datasets, with no clear monotonic pattern concerning the missing rate.

**Figure 1:**
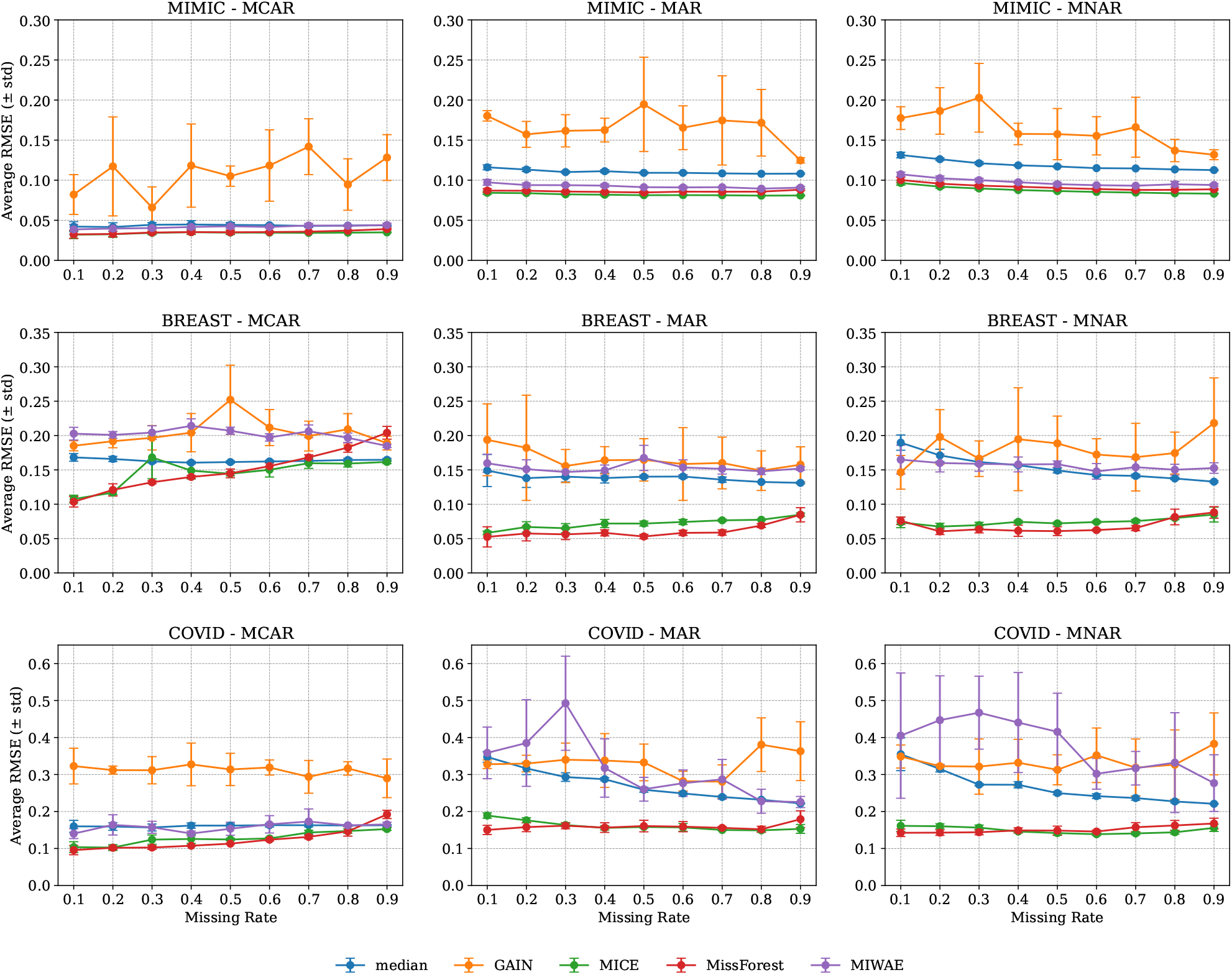
RMSE values of five imputation methods across missing rates (10% to 90%) under MCAR, MAR, and MNAR mechanisms in the MIMIC-III, Breast Cancer, and COVID-19 datasets.

### Deep learning-based imputation methods show greater variability in AUC

As shown in Fig 2, when the missing rate is low, all imputation methods achieve relatively similar AUC scores in downstream classification tasks. In most cases, MICE and MissForest slightly outperform deep learning-based methods such as GAIN and MIWAE.

**Figure 2:**
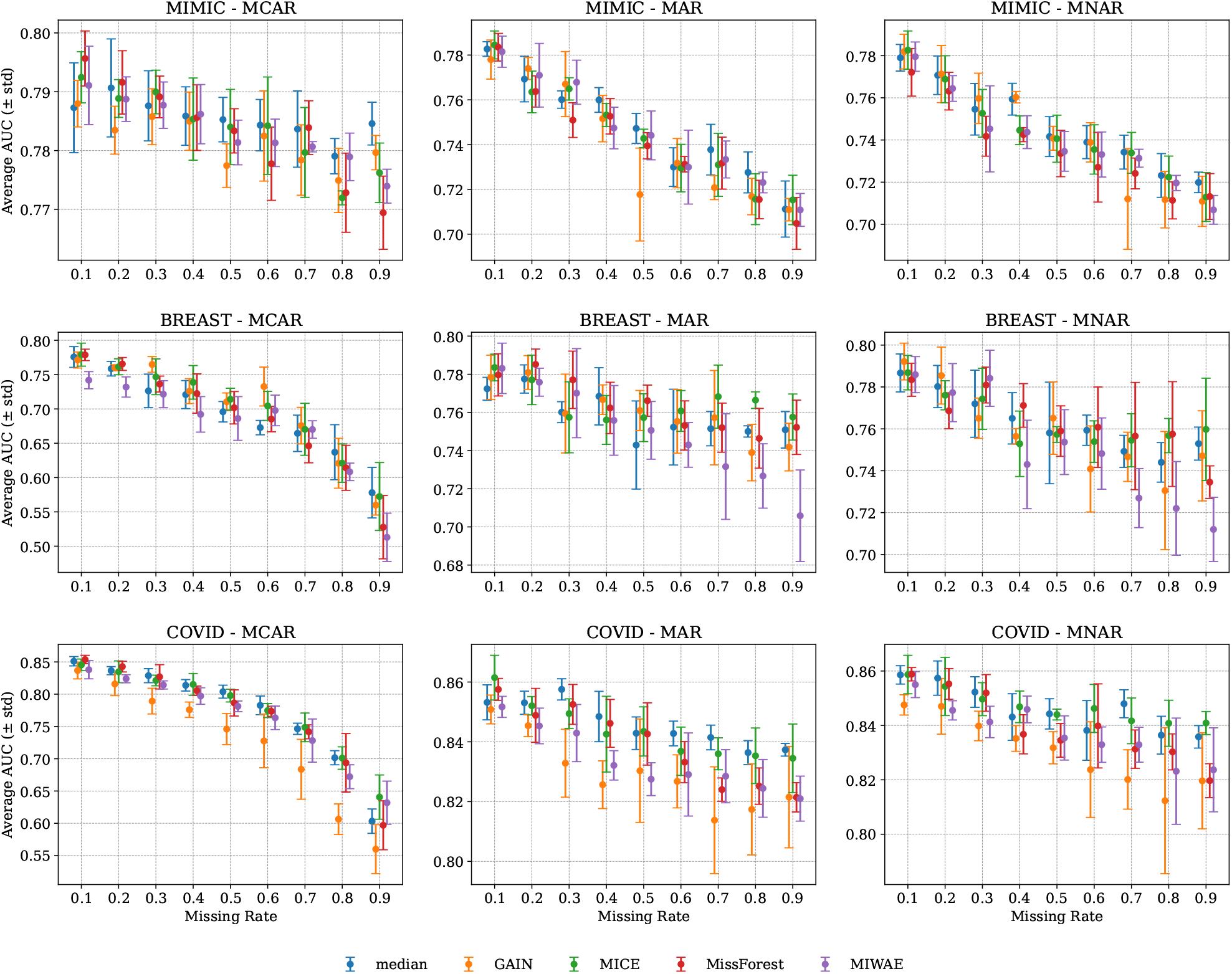
AUC values of five imputation methods across missing rates (10% to 90%) under MCAR, MAR, and MNAR mechanisms in the MIMIC-III, Breast Cancer, and COVID-19 datasets.

As the missing rate increases, the AUC performance of different imputation methods begins to diverge more noticeably. Under the MNAR mechanism, GAIN and MIWAE exhibit greater variability in AUC, while traditional methods such as MICE and MissForest demonstrate more consistent performance.

On the Breast Cancer dataset, MIWAE yields the lowest AUC values among all methods across most settings. The largest differences are observed under the MAR and MNAR mechanisms at 50% missingness. In contrast, MICE exhibits relatively stable AUC values across all mechanisms and missingness levels.

AUC values varied depending on the location of missingness, indicating that the impact of missing data is not uniformly distributed across all features (see Supplementary Figure S1), and that the position of missing values can differentially affect downstream model performance.

Across all mechanisms, an increase in the missingness rate generally leads to a decrease in AUC. This trend is particularly pronounced under the MCAR mechanism. In contrast, the decline in AUC is more gradual under MAR and MNAR settings.

### Inconsistency analysis of imputation effect and predictive performance

We compared the ranking of five imputation methods based on both RMSE and AUC across varying missing rates and mechanisms, as shown in Figure 3. Rankings were assigned from 1 (best) to 5 (worst), with lower values indicating better performance. Interestingly, the method that achieves better rankings in RMSE does not necessarily yield better AUC in classification tasks. For example, MICE frequently ranks best in RMSE across multiple missing mechanisms and rates, but its AUC rankings vary significantly and are not always among the top. In contrast, GAIN, despite often showing the worst RMSE rankings, attains relatively better AUC under MAR and MNAR conditions, especially when the missing rate is below 70%.

**Figure 3:**
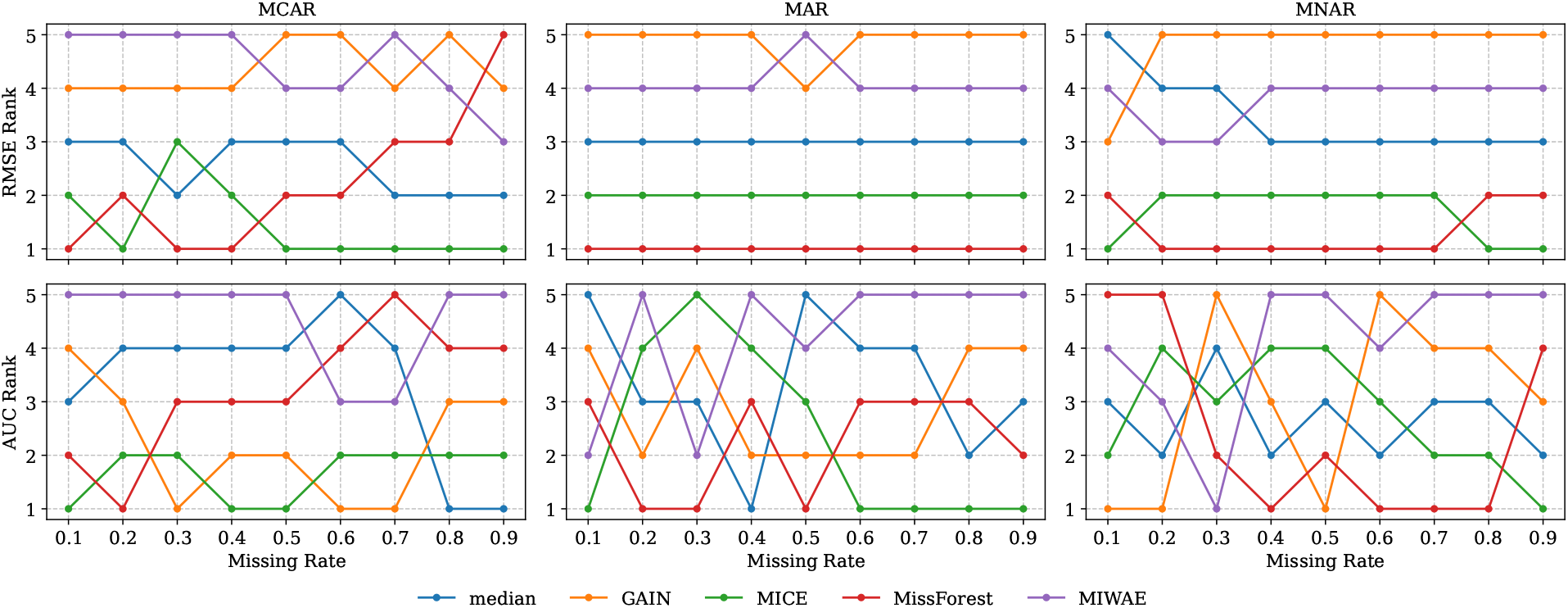
The RMSE and the AUC of the five methods in Breast Cancer were ranked relative to each other, with smaller numbers indicating better results.

### Traditional methods maintain more stable performance across mechanisms and datasets

To assess the comparative performance of imputation methods, we computed the difference between each method’s score and the mean score across all methods within the same setting (i.e., dataset, missingness mechanism, and missingness rate). Figure 4 visualizes these deviations for RMSE and AUC across three datasets (MIMIC-III, Breast Cancer, and COVID-19), under MCAR, MAR, and MNAR mechanisms at 20% and 50% missingness. Each point represents the average deviation over five runs per setting. In the RMSE plots (top row), methods located farther to the right indicate larger RMSE values compared to the average, and hence poorer reconstruction accuracy. Across most settings, MICE and MissForest appear on the left side, indicating relatively lower RMSE. In contrast, Median consistently appears on the right side across mechanisms and datasets, indicating larger reconstruction errors. GAIN and MIWAE also exhibit higher RMSE under some conditions, with greater spread observed at 50% missingness.

**Figure 4:**
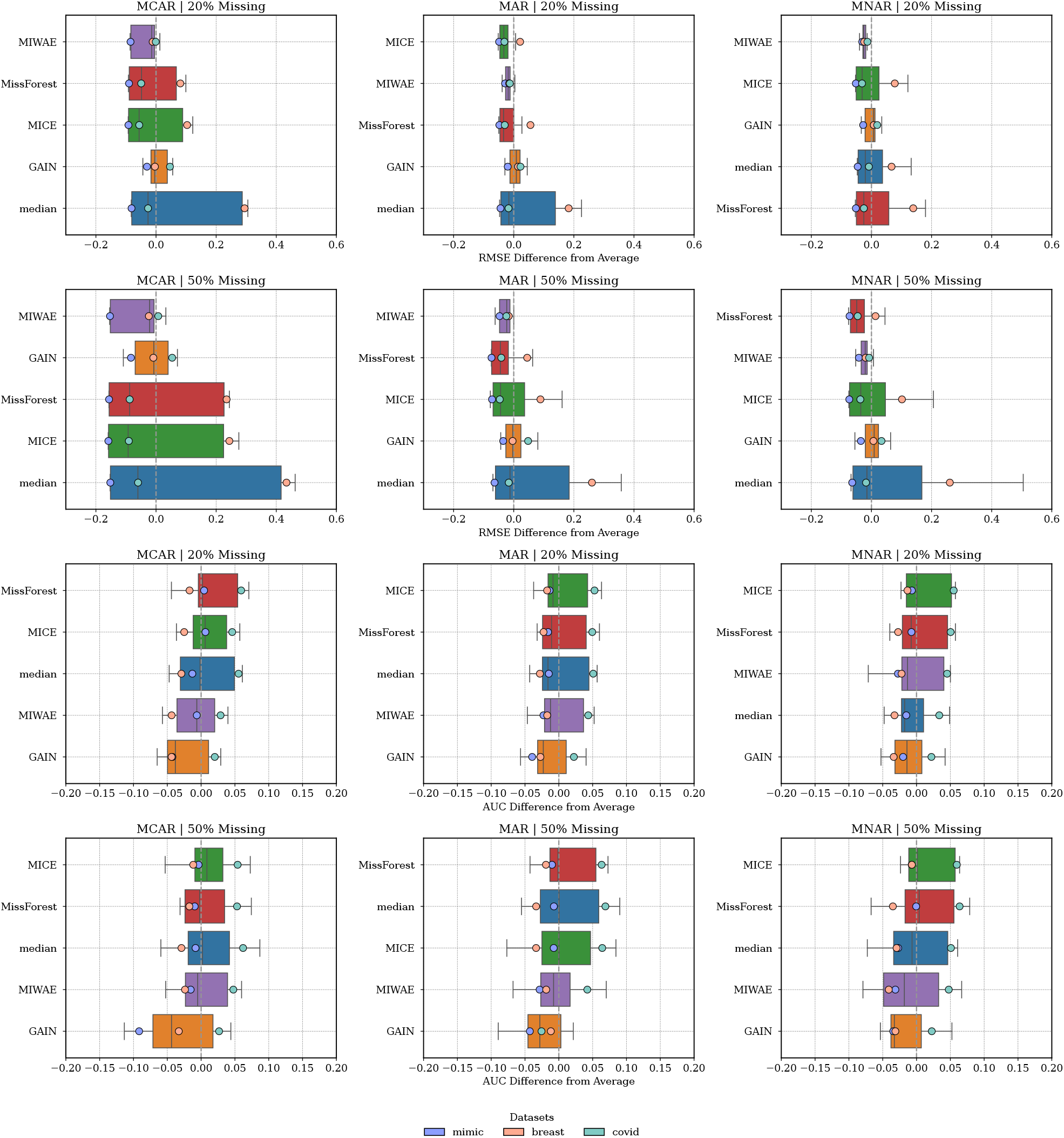
Mean deviation of RMSE and AUC for each imputation method across three datasets (MIMIC-III, Breast Cancer, and COVID-19) under MCAR, MAR, and MNAR mechanisms at 20% and 50% missingness. Each point represents a dataset, with the horizontal axis indicating the difference between the method’s average RMSE (or AUC) and the mean of all methods under the same setting. Points to the left/right reflect worse/better performance relative to the average. This figure illustrates the relative effectiveness of each method under different missingness conditions.

In the AUC plots (bottom row), methods positioned to the right indicate better classification performance relative to the average. MICE and MissForest are generally close to or slightly above the mean in most settings. GAIN appears on the left side in all conditions, indicating AUC values consistently below the average. MIWAE varies more across settings and shows greater deviation under MAR and MNAR at higher missing rates. Median shows more variable behavior, with some settings above and others below the average.

Increasing the missingness rate from 20% to 50% generally leads to higher RMSE, reflecting reduced imputation accuracy. However, AUC remains relatively stable for most methods under MCAR and MAR, indicating that classification performance may be preserved even when numeric reconstruction deteriorates, especially when the data structure remains predictable. MICE and MissForest show minimal degradation in both metrics as missingness increases, underscoring their robustness.

Performance disparities are most evident under MCAR, where the randomness of missingness limits information recovery and results in larger variability across methods. Under MAR, performance becomes more consistent as methods can exploit observed-variable relationships. MNAR, the most challenging setting due to unobserved-value dependence, sees performance decline across all methods—but MICE and MissForest still maintain relative advantage.

Dataset characteristics are also associated with differences in imputation performance. On the MIMIC dataset, which contains mostly continuous features, MICE and MissForest show lower RMSE values and higher AUC values in most settings. On the Breast Cancer dataset, where many features are categorical, GAIN and MIWAE yield higher RMSE and lower AUC in several settings, particularly under MAR and MNAR mechanisms. On the COVID-19 dataset, the performance of all methods is more similar, with smaller deviations observed across both metrics.

### Joint visualization reveals that traditional methods consistently balance imputation accuracy and classification performance

To provide an overview of imputation performance under varying missing data conditions, we visualized the results of five commonly used methods across three clinical datasets (Figure 5). Each circle represents a method’s performance under a specific combination of missing mechanism and missing rate. The size of each bubble corresponds to the average RMSE (with smaller bubbles indicating lower imputation error), while the color intensity reflects the average AUC (with warmer colors representing higher classification performance).

**Figure 5:**
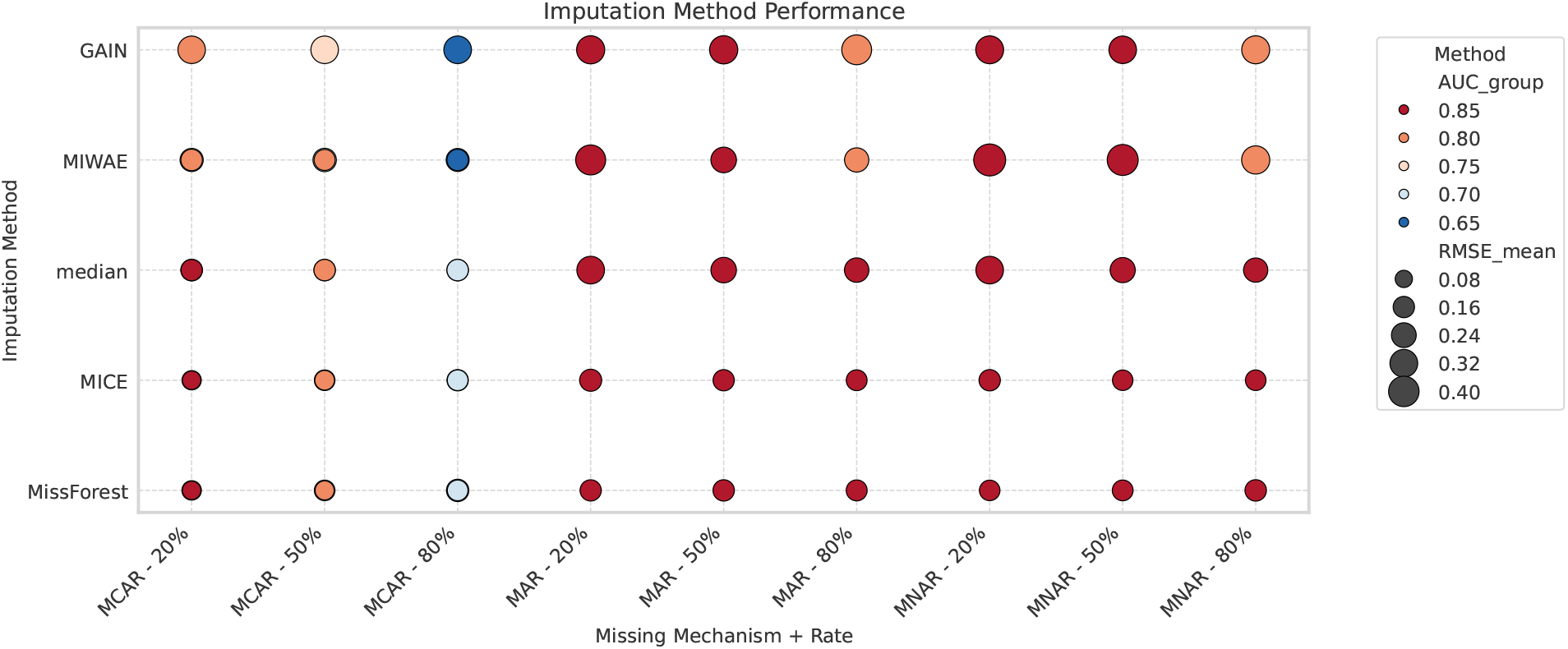
Comparison of five imputation methods across three clinical datasets under different missingness mechanisms and rates. Each circle represents a method’s performance under a specific condition. Bubble size indicates RMSE (larger size = higher error), and color intensity reflects AUC (darker red = better classification performance).

Across the datasets, traditional methods such as MICE and MissForest frequently demonstrated smaller imputation errors and consistently favorable downstream classification performance. In contrast, deep learning–based methods, particularly GAIN and MIWAE, showed more variable results depending on the missing data mechanism and rate. For example, GAIN achieved relatively high AUC under MCAR with low missingness, but its imputation error (RMSE) remained comparatively higher in several settings.

## DISCUSSION

In this study, we systematically evaluated the performance of five widely used imputation methods—Median, MICE, MissForest, GAIN, and MIWAE—across three types of missing data mechanisms (MCAR, MAR, and MNAR) with a range of missing rates in three datasets (MIMIC, Breast Cancer, and COVID-19). Our goal was to understand how different imputation strategies affect both the accuracy of value imputation and the performance of downstream, and to guide the selection of imputation methods under differenct missing scenarios.

Traditional imputation methods, particularly MICE and MissForest, demonstrated more stable and reliable performance under diverse conditions. These methods consistently achieved better rankings in terms of lower root mean squared errors (RMSE) and higher area under the curve (AUC) scores, especially under challenging settings such as MNAR and higher missingness rates. This is consistent with findings from prior evaluations^21,22^, which showed that tree-based and regression-based imputers offer more reliable performance across structured datasets. However, unlike^23^, we find that these methods also scale well under higher missingness and non-random mechanisms.One possible explanation is that MICE and MissForest leverage strong structural assumptions—such as linear or tree-based dependencies between features—which enable them to remain robust even when key information is partially missing.^24–26^Their performance distributions were also more concentrated with lower variance, indicating stronger adaptability across datasets and missingness mechanisms.

In contrast, deep learning-based methods such as GAIN and MIWAE showed greater variability, particularly in their AUC performance under MCAR and high missingness conditions. This observation may be attributed to their reliance on complex neural architectures, which are more sensitive to limited or irregularly missing training data and may struggle to generalize when missingness patterns are unpredictable. While prior studies have reported that GAIN can outperform traditional methods under certain conditions^27,28^, our results suggest that its performance advantage does not consistently hold across datasets or mechanisms, especially in small-sample or highly sparse settings. Our findings suggest that the performance gains reported in synthetic or fully numerical datasets^8^ may not generalize to heterogeneous, clinical-like tabular data. This contrasts with earlier claims of general superiority of deep learning methods for imputation.

A key insight from our ranking analysis is the decoupling between imputation accuracy and downstream task performance. In several cases, methods with lower RMSE did not yield the highest AUC, while those with higher reconstruction error still achieved competitive or superior classification results. This challenges the common assumption that accurate pointwise imputation is sufficient, and instead emphasizes the need to evaluate imputation methods in the context of their downstream utility. Our findings echo and extend those of previous work^22^, which provided a notable and timely critique of conventional imputation evaluation paradigms. Their study convincingly demonstrated that imputation quality does not always translate to improved model performance, a perspective that has significantly influenced the framing of our own experiments. Moreover, while previous studies have noted the disconnect between imputation quality and downstream performance, our use of method-wise ranking across RMSE and AUC offers a more systematic quantification of this divergence. By building on their insights, we aim to provide a broader empirical foundation to better understand when and why such mismatches occur.

Moreover, we observed that missingness location played a significant role. When high-importance features were missing, the degradation in classification performance was more substantial than when less informative features were affected—even when overall RMSE remained moderate. This heterogeneous effect suggests that imputation errors in critical features disproportionately influence model outcomes, and future imputation strategies may benefit from incorporating feature importance weighting. This effect was particularly pronounced for GAIN and MIWAE, further illustrating their sensitivity to data structure.

Performance also varied across datasets. On the MIMIC dataset, which primarily comprises continuous variables, traditional methods such as MICE and MissForest performed consistently well in both imputation and classification tasks. In contrast, on the Breast Cancer dataset, which includes a large proportion of categorical features, MIWAE exhibited lower and more variable AUC scores, particularly under MAR and MNAR mechanisms. While MIWAE has shown strong performance in prior studies involving numerical datasets with low-dimensional latent spaces^9^, our results suggest that its effectiveness may diminish when applied to datasets with high categorical proportion and limited training samples.This underperformance may be attributed to its limited capacity to effectively model structured categorical data. For the COVID-19 dataset, method differences were generally less pronounced; however, MissForest demonstrated the most stable performance across all mechanisms. These findings highlight the importance of considering dataset characteristics—such as variable types and structure—when selecting imputation methods, and suggest that deep learning-based imputers may face challenges in generalizing to heterogeneous or tabular datasets.

From the perspective of missingness mechanisms, MCAR yielded the most pronounced separation in method rankings, likely due to the lack of structured missingness patterns which certain models, especially deep learning-based ones, may implicitly rely on. Although MNAR is often considered the most challenging mechanism due to unobserved dependencies^29^, our results reveal that deep models can also exhibit high variability under MCAR. This contrasts with assumptions in some prior evaluations that treat MCAR as a relatively benign baseline.This suggests that instability can stem not only from the complexity of missingness patterns but also from the randomness and lack of signal in missing locations.

While recent studies have promoted deep learning-based imputation models for their flexibility and theoretical advantages, our empirical findings suggest that traditional methods remain highly effective in structured clinical datasets. The frequent appearance of MICE and MissForest as top-ranked methods highlights their robustness across mechanisms and missingness levels. Importantly, these methods offer reliable performance with lower computational complexity and greater interpretability—features that are valuable in clinical settings. Despite the overall strength of traditional methods, deep learning models such as GAIN showed promising results in specific settings. For instance, in the COVID-19 dataset, GAIN achieved competitive AUC scores under MCAR at high missingness levels. This suggests that deep models may be better suited for datasets with less heterogeneous variable types or more stable feature dependencies.

## Conclusion

In this study, we systematically evaluated five representative imputation methods—Median, MICE, MissForest, GAIN, and MIWAE—across three real-world clinical datasets (MIMIC, Breast Cancer, and COVID-19) under varying missingness mechanisms (MCAR, MAR, and MNAR) and missing rates (10% to 90%). We assessed both imputation accuracy (RMSE) and downstream classification performance (AUC), providing a comprehensive comparison of traditional and deep learning-based imputation strategies.

Our findings suggest that traditional methods,especially MICE and MissForest,remain highly robust and effective in clinical datasets with structured tabular formats. These methods consistently achieved top rankings across metrics and settings, and exhibited lower performance variance, particularly under challenging missingness patterns such as MNAR or high missing rates. In contrast, deep learning-based methods like GAIN and MIWAE showed greater variability and were more sensitive to the data structure and missingness mechanisms. While GAIN occasionally achieved competitive performance under favorable conditions, its results were less stable across settings.

Furthermore, our ranking-based analysis revealed a notable divergence between imputation accuracy and downstream predictive performance, as reflected in the inconsistent alignment between RMSE and AUC rankings across methods and settings. This finding aligns with prior theoretical and empirical work^30^ suggesting that improvements in imputation quality often yield only marginal gains in prediction performance—especially when using flexible models or under uninformative missingness mechanisms. Our empirical evidence extends these insights to heterogeneous, real-world datasets with mixed data types, emphasizing the need to evaluate imputation strategies not only by reconstruction error, but also by their impact on the specific predictive tasks of interest.

Rather than prescribing a fixed imputation strategy, our findings aim to illuminate how method performance varies under different data and missingness characteristics. By systematically comparing imputation quality and downstream utility across multiple conditions, we provide empirical evidence that may help researchers better understand the trade-offs involved in imputation. These observations underscore the importance of considering data structure, missingness mechanisms, and modeling objectives jointly, especially when imputation is used as a pre-processing step in real-world clinical modeling pipelines.

## Data Availability

The MIMIC-III dataset is available via PhysioNet (\url{https://physionet.org/content/mimiciii/1.4/}) after completing the required training and data use agreement. In this study, we used a preprocessed version of the MIMIC-III dataset provided by \citep{shadbahr2023impact} as part of their published work.The Breast Cancer dataset used in this study is a processed subset originally derived from data at Memorial Sloan Kettering Cancer Center (MSKCC), made available by \citep{shadbahr2023impact}. The original raw data are not publicly available due to patient privacy, but we used the processed version provided by the authors for research use.The COVID-19 dataset used in this study is publicly available from Kaggle at: \url{https://www.kaggle.com/datasets/S%C3%ADrio-Libanes/covid19/data}. Access requires a Kaggle account and agreement to the dataset’s terms of use.

## Supplementary Information

**S1 Fig**. *AUC Distribution under Different Missing Variable Settings*. As an illustrative example, this figure shows the variation in AUC values under the MAR mechanism in the Breast Cancer dataset, where different columns were randomly selected as the missing variable.

## RESOURCE AVAILABILITY

### Data Availability

The MIMIC-III dataset is available via PhysioNet (https://physionet.org/content/mimiciii/1.4/) after completing the required training and data use agreement. In this study, we used a preprocessed version of the MIMIC-III dataset provided by^22^ as part of their published work.The Breast Cancer dataset used in this study is a processed subset originally derived from data at Memorial Sloan Kettering Cancer Center (MSKCC), made available by^22^. The original raw data are not publicly available due to patient privacy, but we used the processed version provided by the authors for research use.The COVID-19 dataset used in this study is publicly available from Kaggle at: https://www.kaggle.com/datasets/S%C3%ADrio-Libanes/covid19/data. Access requires a Kaggle account and agreement to the dataset’s terms of use.

### Code Availability

The code will be made publicly available upon publication at: https:.

